# Oronasal mucosal melanoma is defined by two transcriptional subtypes in humans and dogs with implications for diagnosis and therapy

**DOI:** 10.1101/2024.10.18.24315503

**Authors:** Kelly L. Bowlt Blacklock, Kevin Donnelly, Yuting Lu, Jorge del Pozo, Laura Glendinning, Gerry Polton, Laura Selmic, Jean-Benoir Tanis, David Killick, Maciej Parys, Joanna Morris, Inge Breathnach, Stefano Zago, Sara M Gould, Darren Shaw, Mickey Tivers, Davide Malucelli, Ana Marques, Katarzyna Purzycka, Matteo Cantatore, Marie E. Mathers, Mark Stares, Alison Meynert, E. Elizabeth Patton

**Affiliations:** Royal (Dick) School of Veterinary Studies and the Roslin Institute, Edinburgh, EH25 9RG, UK; MRC Human Genetics Unit, Institute of Genetics and Cancer, University of Edinburgh, Edinburgh EH4 2XU, UK; CRUK Edinburgh Centre, Institute of Genetics and Cancer, University of Edinburgh, Edinburgh EH4 2XU, UK; North Downs Specialist Referrals, 3&4 The Brewerstreet Dairy Business Park, Brewer Street, Bletchingley, RH1 4QP, UK; Department of Veterinary Clinical Sciences, The Ohio State University, 601 Vernon L. Tharp St., Columbus, OH 43210, USA; School of Biodiversity, One Health and Veterinary Medicine, University of Liverpool. Leahurst Campus, Chester High Road, Neston, Wirral, CH64 7TE, UK; University of Glasgow, Bearsden Road, Glasgow G61 1QH, UK; The Ralph Veterinary Referral Centre, Fourth Avenue, Globe Business Park, Marlow, SL7 1YG, UK; University of Bristol, Langford, Bristol, BS40 5DU, UK; Paragon Veterinary Referrals, Paragon Point, Red Hall Crescent, Wakefield, WF1 2DF, UK; VetsNow, 123-145 North Street, Glasgow, G3 7DA, UK; Anderson Moores Veterinary Specialists, The Granary, Bunstead Barns, Hampshire, SO21 2LL, UK; Department of Pathology, Western General Hospital, Crewe Road, Edinburgh, EH4 2XU, UK; Edinburgh Cancer Centre, Western General Hospital, Crewe Road, Edinburgh, EH4 2XU, UK

**Keywords:** Mucosal melanoma, Canine, Human, Oronasal, Transcriptome, CTLA4, cMET, Immunity, Microbiome, Macrophages

## Abstract

Mucosal melanoma is a rare melanoma subtype associated with a poor prognosis and limited existing therapeutic interventions, in part due to a lack of actionable targets and translational animal models for pre-clinical trials. Comprehensive data on this tumour type is scarce, and existing data often overlooks the importance of the anatomical site of origin. We evaluated human and canine oronasal mucosal melanoma to determine whether the common canine disease could inform the rare human equivalent.

Using a human and canine primary oronasal mucosal melanoma (OMM) cohort of treatment naive archival tissue, alongside clinicopathological data, we obtained transcriptomic immunohistochemical, and microbiome data from both species. We defined the transcriptomic landscape in both species, and linked our findings to immunohistochemical, microbiome and clinical data.

Human and dog OMM stratified into two distinctive transcriptional groups which we defined using a species-independent 41-gene signature. These two subgroups are termed CTLA4-high and cMET-high, and indicate actionable targets for OMM patients. To guide clinical decision-making, we developed immunohistochemical diagnostic tools which distinguish between transcriptomic subgroups.

For the first time, we find that OMM has conserved transcriptomic subtypes and biological similarity between the canine and human OMM, with significant implications for patient classification, treatment, and clinical trial design.

## Introduction

Mucosal melanoma (MM) is a rare and aggressive form of melanoma arising from melanocytes in sun-protected mucosal surfaces, including oral, nasal, genital, and anorectal regions [1]. Despite accounting for a minority (1.4%) of melanoma cases, MM carries a disproportionately higher morbidity and mortality risk than cutaneous melanoma (CM) [2]. MM is often diagnosed at an advanced stage, exhibits a higher likelihood of metastasis (44% in MM; 14% in CM) [3,4], and a reduced 5-year survival (up to 25-31% in MM; 93% in CM) [3,5–7]. Unlike CM, MM lacks well-defined precursor lesions, and has few oncogenic driver mutations or effective therapeutics[2,3,8–10]. MM from different anatomical sites is mutationally heterogeneous, although interestingly, there is some evidence for “upper” and “lower” body site-specific mutational profiles [4,11].

We reasoned that exploring the nuances of anatomical site-specific MM, with a specific focus on oronasal mucosal melanoma (OMM), has the potential to disentangle the inherent heterogeneity of this rare melanoma subtype. There are few experimental models for OMM [12–14], however OMM is a common and naturally occurring oral tumour in dogs. Previous research has identified two molecular subgroups in OMM in dogs based on RNA expression, but the clinical significance of these subgroups for veterinary oncology is unknown [15]. Despite dogs being proposed as a model for human OMM, it is unknown if these subtypes even exist in human OMM, and whether human OMM is analogous to the canine disease. These outstanding questions are critical because the lack of diagnostic subtypes precludes subtype classification that could be pivotal for patient-centric precision medicine and could inform clinical trial design to account for potential subtype-specific responses [9,16–21].

Here, for the first time, we present transcriptomic and 16s sequencing data for human and canine OMM side by side. Our analysis reveals OMM tumour stratification for both human and canine patients based on a CTLA4-high or cMET-high transcriptional signature, defined by 41 genes, shared between species. This patient stratification is facilitated by an immunohistochemical tool, pivotal not only for individualised patient care but also for informing clinical trial design.

## Results

### OMM in human and dogs share clinicopathological characteristics and poor prognosis

We identified human (Edinburgh cohort; n=17) and canine (Bowlt Blacklock cohort; n=36) patients who presented for treatment of naturally occurring, treatment-native OMM and had archival formalin-fixed-paraffin-embedded (FFPE) tissue and clinical records available for analysis (**Figure 1A**). In both humans and dogs, OMM can appear as melanotic or amelanotic, and are most frequently observed in the oronasal cavity in humans and in the oral cavity in dogs (**Figure 1B**). In both species, underlying bone lysis secondary to local tumour invasion is common (**Figure 1C**). Histopathological findings are also similar, consisting of proliferations of melanotic or amelanotic infiltrates of pleomorphic, round and/or spindloid neoplastic cells that invade both the lamina propria and deep connective tissue (**Figure 1D**). RNA sequencing and 16s sequencing data were assimilated and compared to clinical data. When considering survival, the time from OMM diagnosis to end-of-life is not directly comparable between species. Human patients die of their disease, whereas in dogs, 89.3% of recorded deaths are a result of euthanasia [22,23].

**Figure 1:**
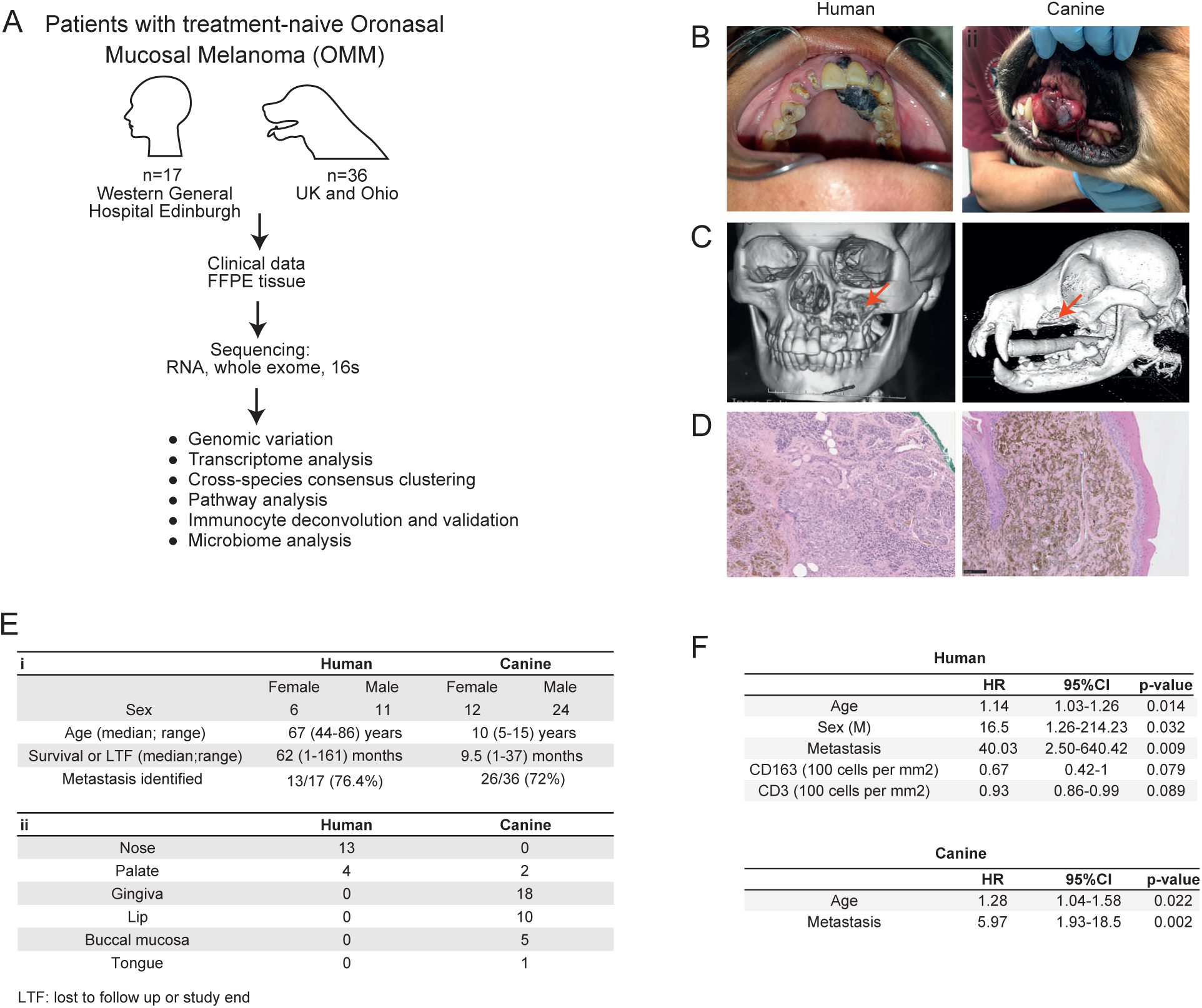
OMM in human and dogs share clinicopathological characteristics and poor prognosis. **A**: Project workflow. **B:** Gross appearance of human and canine primary OMM. **C**: 3D computed tomography (CT) reconstruction images showing left sided maxillary destruction (red arrow) underlying the primary tumour in a human and canine patient. **D:** Haematoxylin and eosin staining showing that both human and canine OMM exhibits similar proliferations of pigmented neoplastic infiltrates. **E:** Overview of clinical pathological data for the study cohort, encompassing (i) patient data, and (ii) anatomical location of the primary OMM. **F**: Relevant hazard ratios for survival in human and canine patients with OMM based on clinical parameters and intra-tumoural immunocyte infiltration. *Images of human patients (*Figure 1A and B*) are kindly provided by Professor Ankita Chugh BDS MDS, All India Institute of Medical Sciences Jodhpur (copyright retained)*.

We investigated the relationships between clinicopathological features, treatment, and survival. Clinicopathological data is presented in **Figure 1E**. The best model to identify predictors of survival in humans included age, gender (male), presence of metastasis, and immunocyte infiltration (**Figure 1F**). Risk of death increased by 14% for every one-year increase in age, 16.5-fold for male patients, and 40-fold for patients with metastatic disease (**Figure 1F**). In dogs, the best model to identify predictors of survival included only age and presence of metastasis (**Figure 1F**). Human patients who only underwent biopsy of their OMM without surgery had a significantly increased risk of death compared to those who received curative-intent surgery **(Supplementary Figure 1A)**. We found no significant correlation between the relative hazard of death and provision of chemotherapy, radiation therapy or immunotherapy in humans or dogs **(Supplementary Figure 1A)**. In our canine patients, reduced survival probability was identified in patients with WHO stage 3-4 disease, and ulcerated tumours **(Supplementary Figure 1B,C**). Exophytic status, melanotic status and melanoma histopathological subtype were not associated with survival in dogs **(Supplementary Figure 1D-F)**. Equivalent data were not available for human patients.

In conclusion, for both human and canine patients with OMM, our clinical data reveals that prognosis is poor, and metastasis is common. In humans, our data shows that curative surgery and being female is associated with improved survival, while radiation, immunotherapy, and chemotherapy do not significantly impact survival in either species.

### OMM can be stratified into two transcriptomic subgroups

Despite no widespread actionable mutation targets in OMM, we hypothesized that OMM may have transcriptional subtypes, as found in cutaneous melanoma [24]. We conducted RNA-seq transcriptomic analysis of OMM from human (Edinburgh) and canine (Bowlt Blacklock) patients, and performed unsupervised hierarchical clustering from these cohorts as well as from a published canine OMM RNA dataset [15]. Critically, we identified two stable consensus clusters (A and B) in both species using optimal consensus partitioning method, using median absolute deviation and spherical k-means clustering (**Figure 2A-C**). Principal component analysis showed minimal overlap between these transcriptional subgroups in all three datasets **(Supplementary Figure 2A)**. To understand the pathways that characterised the transcriptional subtypes, we applied over-representation analysis (ORA) and showed that KEGG pathways and biological processes align with an immune response in consensus cluster group B (**Figure 2D**). In contrast, ORA of consensus cluster group A was less clear but highlighted pathways associated with the cell cycle and DNA/RNA processing and repair **(Supplementary Data 1)**.

**Figure 2:**
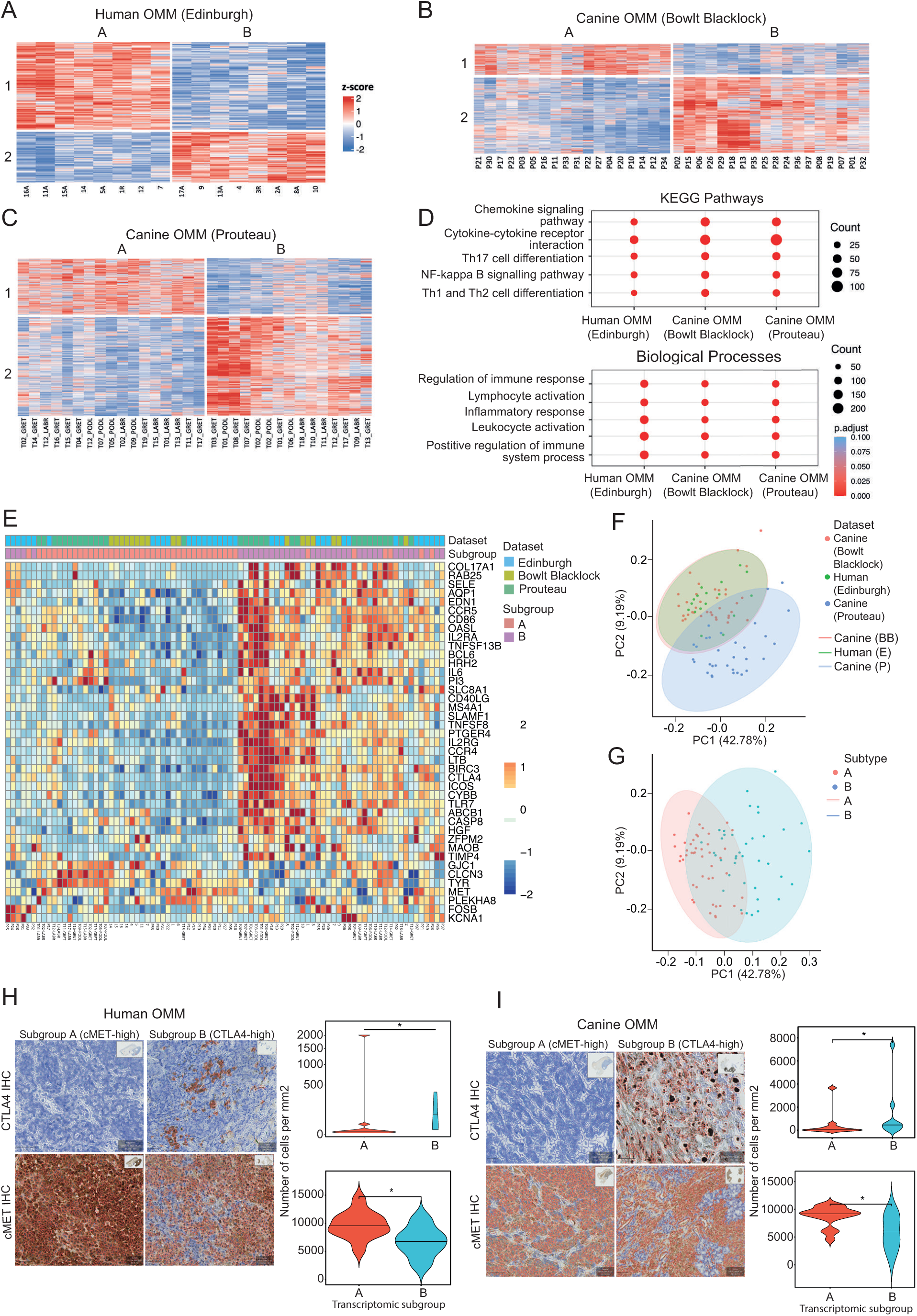
OMM can be stratified into two transcriptomic subgroups. **A-C:** Full length view of the unsupervised cluster diagrams performed using ‘cola’ and MAD:skmeans on bulk RNA sequencing data obtained from OMM from human (Edinburgh; n=17 patients), canine (Bowlt Blacklock; n=36 patients) and canine (Prouteau; n=32 patients) patients. Patients are orientated along the vertical axis and genes are orientated along the horizontal axis. **D:** Group B aligned with the immune response in both KEGG pathways and biological processes**. E:** Heatmap produced using a 41-gene signature to stratify patients into transcriptomic subgroups, independent of species. **F-G:** Principal component analysis (PCA) plot of 812 homologous genes, stratified according to (**F**) species and (**G**) transcriptomic subgroup**. H-I:** Violin plots showing cMET and CTLA4 immunohistochemistry markers in (**H**) human, and (**I**) canine OMM according to transcriptomic subgroup.

To investigate whether the transcriptional subgroups shared between human and canine OMM cohorts were mediated through overlapping gene networks, we aimed to find the most important variables that drive the stratification across species and cohorts. We used the machine learning algorithm randomForest [25] and the 812 human and canine homologous genes (**Supplementary Data 2)**. Across five randomly selected seeds, this approach generated successful models (trained on 50% of the samples) that predicted the B subgroup versus the A subgroup in the test data (the remaining 50% of the samples), independent of species (accuracy 79-88%, binomial test, p-value <0.001). Within these models, 41 genes were consistently ranked among the top 100 genes shared by at least four models and 17 genes were shared by all five models (**Supplementary Figure 2B; Supplementary Data 2)**. Using these 41 genes, optimal discrimination between transcriptomic subgroups was achieved by Euclidean distance clustering (**Figure 2E**). Using PCA clustering based on these 41 genes, we found that our human OMM and dog OMM RNA sequencing data closely clustered on PCA, indicating strong similarity (**Figure 2F**). In contrast, the Prouteau dataset demonstrated mildly distinct distribution on the PC2 (9.19% variation) but not PC1, with the latter strikingly driven by the transcriptional subgroup variation (42.78%) (**Figure 2G**). Therefore, we conclude that the PC2 difference is mainly caused by technical variations during cohort establishment (Bowlt Blacklock *versus* Prouteau), while biologically the two OMM transcriptional subtypes did not exhibit any species-dependent variations.

Notably, our signature highlighted CTLA-4 gene expression in both human and dog OMM. This finding was validated by immunohistochemistry (IHC), showing a significantly greater number of CTLA4-positive cells in the B transcriptomic subgroup (human: p=0.01; canine: p=0.03) (**Figure 2H**). We therefore termed this subgroup “CTLA4-high”. Of particular interest in the A subgroup is a small, enriched panel of genes (*GJC1, CLCN3, TYR, MET, PLEKHAB, FOSB, KCNA1*) highlighted in the 41-gene signature. Using g:Profiler, we mapped theses gene functional information sources to detect statistically significantly enriched terms, which included the hepatocyte growth factor receptor (HGFR, also known as cMET) activity pathway and the cMET complex. We performed cMET IHC and found cMET protein levels to be significantly increased in subgroup A in both humans and dog (human: p=0.04; canine: p=0.02) (**Figure 2I**). Therefore, we termed the A subgroup "cMET-high”. Additionally, we noticed that the enriched panel of genes were MITF target genes and suggest the gene expression signatures of this subgroup may be mediated by MITF [26] **(Supplementary Figure 2C)**.

In conclusion, we show that human and dog OMM can be stratified into two distinctive transcriptional groups, termed CTLA4-high and cMET-high, using the same species-independent 41-gene signature, providing actionable targets and highlighting the biological similarity of this disease between the two species.

### CTLA4 and macrophages are diagnostic biomarkers for the OMM transcriptomic subtypes

The presence of potential drug targets, CTLA4 and cMET, within the subtypes indicated that stratification of patients into each subtype could inform therapeutic decision-making and clinical trial design. Because transcriptomic analysis is currently not practical in most clinical settings, we attempted to identifying diagnostic IHC markers to discriminate between subtypes.

We reasoned that the CTLA4-high subtype could be identified by immunocyte infiltrate populations and utilised the computational framework CIBERSORTx[27] to discern the composition of tumour-infiltrating immune cells from bulk RNA-sequencing data[27] **(Supplementary Data 3).** In both humans and dogs, total immune cell infiltration (the “Absolute Score”) was significantly larger in the CTLA4-high transcriptomic subgroup (p<0.01 in both species), as well as the total B cells, monocyte/macrophages, and T cells (**Supplementary Figure 3A-C**).

Next, to extrapolate these data to a diagnostic test, we used IHC and digital pathology image analysis for immune infiltrate[28]. Utilising species-specific IHC for B cells, monocytes/macrophages and T cells, we observed a higher density of B cells in dogs and higher densities of monocytes/macrophages and T cells in both species in tumour tissues of the CTLA4-high transcriptomic cohort (**Figure 3A-D**). Then, we utilised Receiver Operator Curve (ROC) analysis to determine how well these variables can discriminate between the two transcriptomic subgroups with clinically relevant sensitivity and specificity. Using the Youden index, we identified optimal cut-off values for parameters with an area under the curve (AUC) of >0.75. ROC curve analysis demonstrated that monocytes and macrophages could differentiate between transcriptomic subgroups with high sensitivity in both humans and dogs and with high specificity in humans. This indicates that IHC using CD68 and IBA1 antibodies detecting monocyte and macrophages could be used to inform clinical decision-making based on transcriptomic subtype. In addition, CTLA4 itself (but not cMET) was associated with a clinically relevant sensitivity in both species and a clinically relevant specificity in humans (**Figure 3E**; **Supplementary Figure 3D).**

**Figure 3:**
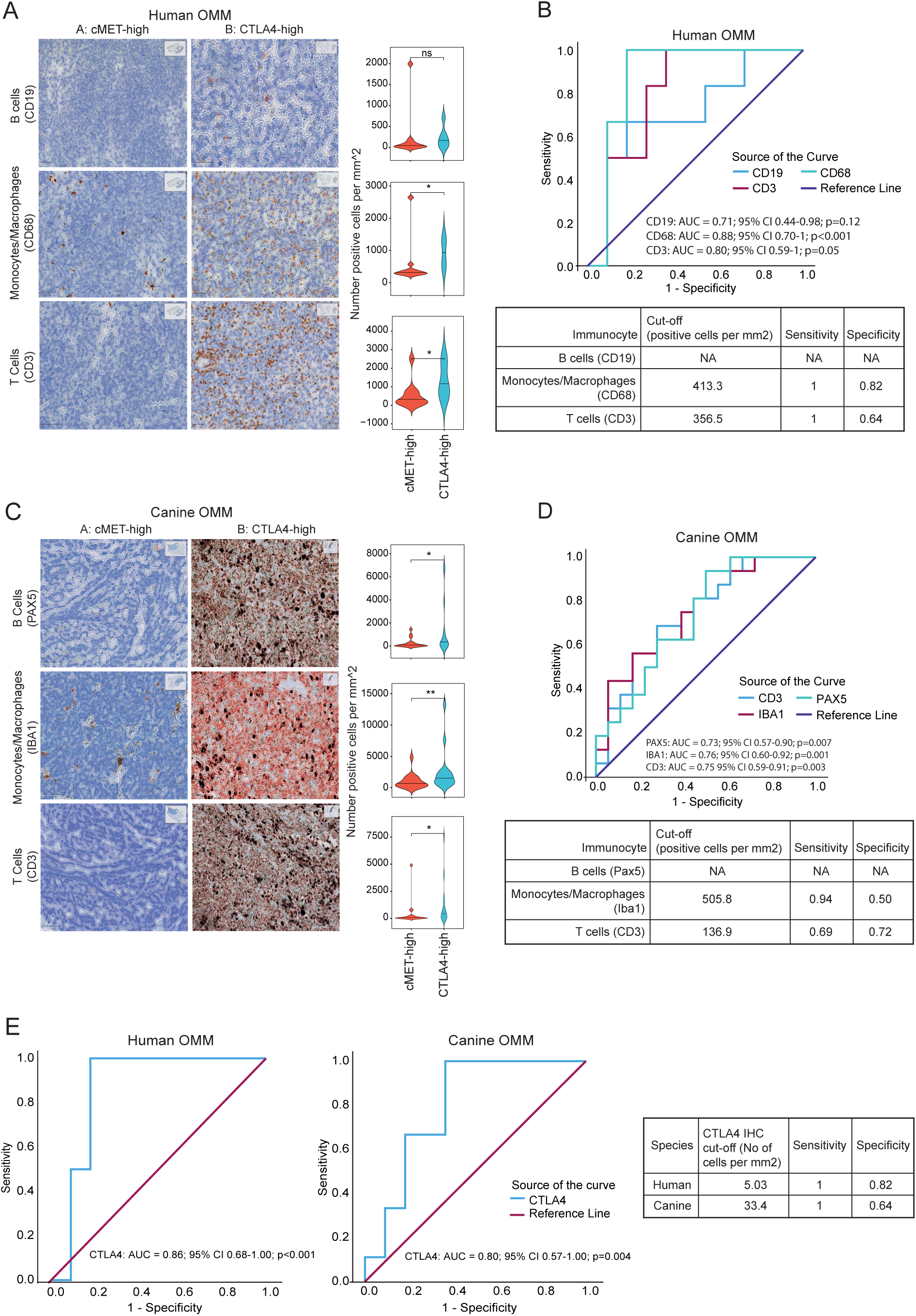
CTLA4 and macrophages are diagnostic biomarkers for the OMM transcriptomic subtypes. **A**: Violin plots and representative IHC panels for human OMM, stratified according to transcriptomic subgroup. **B**: ROC curves were plotted for each immunocyte IHC marker in **A** and optimal cut-off parameters were defined. **C**: Violin plots and representative IHC panels for canine OMM, stratified according to transcriptomic subgroup. **D**: ROC curves were plotted for each immunocyte IHC marker in **C** and optimal cut-off parameters were defined. **E**: ROC curves and optimal cut-off parameters for CTLA4 in human and canine OMM. * p <0.05; **<0.01 ns: not significant

Based on these findings and our RNA-seq analysis, we suggest that the CTLA4-high subtype OMMs are characterised as immune “hot” with high levels of T cell infiltration and activation, and therefore provision of anti-CTLA4 may show favourable response in this subgroup[29]. In contrast, our data suggests the cMET-high tumours are immune "cold" and these patients may have a better response to targeted therapy.

### Tumour microbiota does not correlate with transcriptomic subgroup but may contribute to other pathological features of OMM

The oral cavity hosts the largest and most diverse microbiota after the gut[30]. Given the pivotal role of gastrointestinal tract microbiota in shaping both local and systemic immune system responses[31], we investigated whether the tumour microbiome correlated with our transcriptomic patient stratification, particularly the CTLA4-high transcriptomic subgroup due to the high immune cell infiltration. To this end, we performed 16s sequencing on DNA extracted from FFPE cores harvested from our human and canine tumour cohorts as well as four normal canine oral tissues. Our analysis revealed a significant difference in alpha diversity between species, reflecting the microbiome’s diversity within individual samples, with humans exhibiting significantly lower alpha diversity compared to dogs (**Figure 4A**, p<0.01). Beta diversity, a measure of the similarity or dissimilarity of groups of microbial communities, was significantly different between the two species (p<0.01; **Figure 4B**; **Supplementary Data 4**). Due to the notable differences in alpha and beta diversity observed between species, we conducted separate analyses of the microbiomes associated with human and canine OMM. No significant difference was observed in the microbiome of human or canine OMM when stratified into transcriptomic subtype (**Figure 4B**). Consequently, the two transcriptomic subgroups of OMM were not attributed to an overall difference in the composition of the oral microbiota. Thus, the immunocyte infiltration that we identified above is likely attributed to inflammation with the primary tumour itself and not bacterial burden.

**Figure 4:**
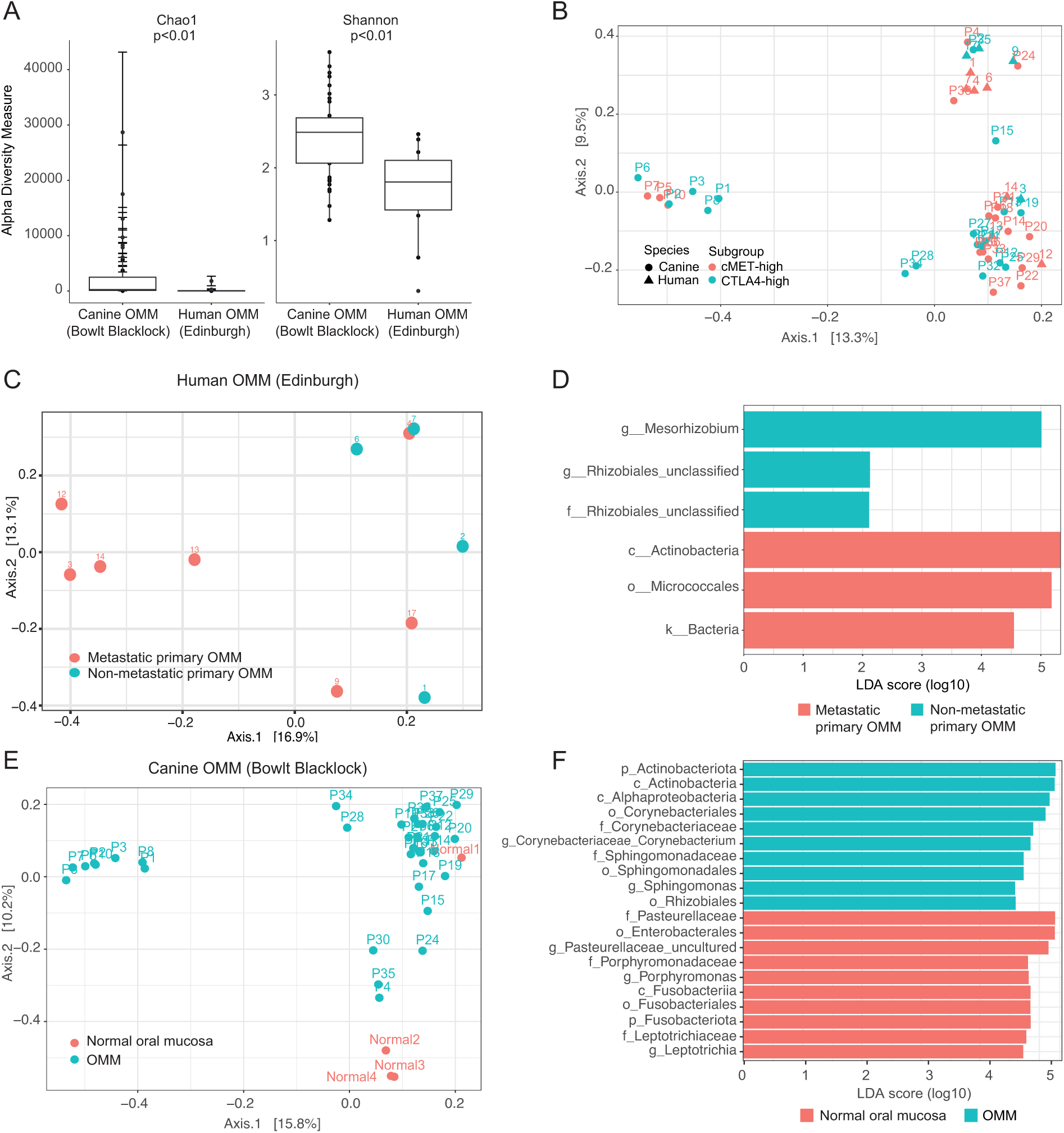
Tumour microbiota does not correlate with transcriptomic subgroup but may contribute to other pathological features of OMM. **A:** Bar and whisker plot of showing the alpha diversity of human and canine OMM. **B**: PCA plot showing the beta diversity of human and canine OMM. **C, D**: In human OMM, overall community diversity is significantly different between metastatic and non-metastatic tumours (p=0.03), with three markers identified that are enriched in each group. **E, F:** In dogs, the overall community diversity is significantly different between tumour and normal oral tissue (p=0.003), with 17 markers enriched in the OMM primary tumour and 111 markers enriched in the normal oral tissue. The top 10 markers in each enriched group are shown.

Although tumour microbiota does not explain the OMM transcriptional subtypes, we did identify two clinically relevant associations that suggest that the tumour microbiome may have a function in OMM disease initiation and progression. First, in human OMM, the overall community diversity was significantly different between metastatic and non-metastatic tumours (beta diversity p=0.03; **Figure 4C**), with three operational taxonomic unit (OTU) markers enriched in each group (**Figure 4D**). This finding suggests that the microbiota composition may influence the metastatic capacity of OMM[32]. Second, we compared the microbiome profiles of canine OMM with those of normal canine oral tissue and found overall community diversity was significantly different between tumour and normal oral tissue (**Figure 4E, F**). These data indicate that OMM correlates with alterations in the oral microbiota composition in dogs, which warrants further investigation because of the emerging role of the microbiome in cancer development and response to therapy[33–35].

## Discussion

Our findings provide the conceptional advance that human and canine OMM is the same disease that can be classified into two subtypes. These two distinct transcriptional subtypes have potential therapeutic implications, and we provide diagnostic tools to distinguish between these two subtypes. Our findings are important for personalised therapeutics for individual patients, and for facilitating patient stratification that may inform clinical trial design and outcomes.

The advent of T-cell targeted immunomodulators, such as Ipilimumab and Nivolumab, and small molecule MAPK-pathway inhibitors has revolutionised cancer care for patients with cutaneous melanoma[36–38]. However, the therapeutic efficacy of immune checkpoint inhibitors for MM remains unclear, and patients have been excluded from most Phase III clinical trials[39]. Therefore, outcome data for patients with MM are scarce and are mainly based on retrospective studies with limited case numbers. Our two transcriptional subtypes suggest that outcomes and clinical trial design could be improved by patient stratification. The CTLA4-high subtype exhibits elevated CTLA4 expression and increased immunocyte infiltration, indicative of a "hot" immune tumour type. Patients with CTLA4-high OMM may benefit from anti-CTLA4 therapy. In contrast, the cMET-high subtype is characterised by high cMET expression, and thus is potentially responsive to small molecule tyrosine kinase inhibitors; the lack of immunocyte infiltration suggests this subtype is a "cold" immune tumour type. Ipilimumab is currently used in clinical patients, but our review of the literature has not identified clinical data regarding ipilmumab or cMET inhibitors as therapeutic approaches for mucosal melanoma. A multicentre retrospective review of treatment efficacy using clinical records would be prudent.

We developed an inexpensive and readily available diagnostic tool, which demonstrates that CTLA4 and monocytes/macrophages distinguish between transcriptomic subgroups with high sensitivity in both humans and dogs. This suggests that IHC utilising CTLA4, CD68 and IBA1 antibodies offer practical utility for guiding clinical decision-making based on transcriptomic profiles.

The extent of immune cell infiltration into tumours has important prognostic value in several cancer types[40–45]. We investigated whether transcriptomic subtype correlated with survival but, unlike in cutaneous melanoma[24,46], this did not reach significance in either human or canine patients. Transcriptional stratification alone may not predict survival because distinct mechanisms contribute to poor prognosis in each subgroup[47,48]. However, notably, we did find that age and metastatic disease was associated with survival in both species. The reason for the survival disparity between genders in our human patients with OMM is unknown. While sex disparities in cancer mortality and survival have been reported previously, particularly for cancers of the floor of the mouth and nasal cavity [49], the specific impact of sex on survival in OMM patients, as observed in our data, remains to be fully understood. Differential environmental exposures and/or physiological processes may explain these disparities[49].

Interestingly, we found no correlation between microbiome diversity and transcriptomic subgroups, suggesting that tumour-associated mechanisms, rather than bacterial infection, drive immune infiltration. However, the observed alterations in oral microbiota associated with OMM development in dogs and OMM metastasis in humans prompts further investigation into the role of microbiota in MM pathogenesis[50–52], although we appreciate that caution is advised when applying sequence-based techniques to the study of microbiota present in low biomass environments[53].

In conclusion, we provide valuable insights into the molecular and immune landscapes of OMM and their potential implications for therapeutic interventions. Our findings are potentially enhanced by the homogeneity of our human cohort, and therefore we underscore the importance of future studies prioritising inclusivity by encompassing diverse cohorts[1,54,55]. Prospective data collection and tissue sampling efforts will augment the database size and mitigate inherent biases and limitations associated with retrospective data quality and completeness. Moreover, future investigations should explore MM from other anatomical sites to determine the generalisability of our findings across different contexts. The implications of our findings suggest that personalised therapeutic strategies targeting shared molecular pathways could hold promise for both human and canine patients with OMM.

## MATERIALS AND METHODS

### EXPERIMENTAL MODEL AND STUDY PARTICIPANT DETAILS

#### Ethics Statements

This study was conducted with the approval of the University of Edinburgh’s Veterinary Ethical Review Committee (Reference number 10.21) and Lothian BioResource Access Committee (REC reference 20/ES/0061 SR1725). Tissue was only included in the study with the informed, written consent of the human patient or the caregiver of the dog who bore the tumour. The treatment that a human or canine patient received was unaffected by their inclusion in this study.

De-identified human clinicopathological and standardised RNA sequencing data have been deposited at the European Genome-Phenome Archive (EGA with accession number #25513).

De-identified canine clinicopathological, genetic and standardised RNA sequencing data have been deposited at the Sequence Read Archive (SRA with accession number #SUB14547907).

De-identified human and canine standardised 16s sequencing data have been deposited at the Sequence Read Archive (SRA with accession number #SUB14547907).

#### Sample collection

Formalin-fixed, paraffin-embedded (FFPE) human and canine OMM tissue samples and associated clinical data were collected from patients with naturally occurring tumours who underwent surgical resection or biopsy of the primary tumour. Tissue was only included in the study with the informed, written consent of the human patient or the caregiver of the dog who bore the tumour.

FFPE tissue samples of treatment-native human OMMs were collected between 2006 and 2021. Human OMM FFPE tissue samples and clinical data were provided by Lothian NRS Bioresource RTB with the approval of Lothian BioResource Access Committee (REC reference 20/ES/0061 SR1725). Clinicopathological data collected included patient gender, ethnicity, age (years), primary tumour location, presence of ulceration, tumour depth (mm) WHO stage, metastatic status, provision of adjuvant therapy, and duration of patient survival or lost to follow up (in months following initial diagnosis of OMM).

FFPE tissue samples of treatment-native canine OMMs were collected between 2011 and 2021 for histopathology from dogs attending the Clinical Veterinary Oncology departments at the Universities of Edinburgh, Bristol, Liverpool, Glasgow and Ohio Small Animal Teaching Hospitals, Vets Now (Glasgow) Referrals, Paragon Referrals, The Ralph Veterinary Referral Centre, Anderson Moores Veterinary Specialists, and North Downs Specialist Referrals. Clinicopathological data collected included sex, breed, age (years), primary tumour anatomical location, melanoma type (melanotic/amelanotic, pleomorphic/composite/nevocytoid/animal, ulcerated, exophytic), depth of invasion (mm), stroma:tumour ratio(%), mitotic count (%), metastatic status (confirmed by abdominal ultrasound, thoracic computed tomography or radiography, and/or histological examination of ≥1 regional lymph node), World Health Organization (WHO) status, provision of adjuvant therapy, and duration of patient survival or lost to follow up (in months following initial diagnosis of OMM).

The diagnosis of OMM was confirmed by a consultant histopathologist (MM) or board certified veterinary anatomic pathologist (JdP), and the regions of the FFPE blocks which represented OMM tissue tumour identified.

## METHOD DETAILS

### RNA and DNA extraction

Using a manual tissue arrayer (Beecher MTA-1, Estigen OÜ, Estonia) machine, two cores (1mm outer diameter, 5mm length) were harvested from each canine FFPE block: one from an area of OMM and one from histopathologically normal tissue. RNA and DNA was isolated from the tissue cores using the Covaris E220 Evolution Focused Ultrasonicator (Covaris Inc, #500429) and the truXTRAC® FFPE total NA (Nucleic Acid) Kit – Column (Covaris, #520220). Inhibitors (e.g. melanin) were removed using the OneStep PCR Inhibitor Removal kit (Zymo Research #D6030).

#### Quality control

DNA (canine only) and total RNA (human and canine) was assessed for quality and integrity on the Fragment Analyser Automated Capillary Electrophoresis System (Agilent Technologies Inc, #5300) with the Genomic DNA 50kb Kit (#DNF-467-0500) and Standard Sensitivity RNA Analysis Kit (#DNF-471-0500) respectively, and then quantified using the Qubit 2.0 Fluorometer (Thermo Fisher Scientific Inc, #Q32866) and the Qubit DNA (# Q32853) or RNA (#Q10210) Broad Range assay kits. In the RNA samples, DNA contamination was quantified using the Qubit dsDNA HS assay kit (#Q32854). The Fragment Analyser data showed a high degree of RNA degradation so the fragmentation step was omitted from the library preparation protocol.

#### RNA library preparation

First-strand cDNA was generated from 50 ng of each total RNA sample using the SMARTer® Stranded Total RNA-Seq Kit v2 – Pico Input Mammalian kit (Clontech Laboratories, Inc. #634411). Illumina-compatible adapters and indexes were then added via 5 cycles of PCR. AMPure XP beads (Beckman Coulter, #A63881) were then used to purify the cDNA library. Depletion of ribosomal cDNA (cDNA fragments originating from highly abundant rRNA molecules) was performed using ZapR v2 and R-probes v2 specific to mammalian ribosomal RNA and human mitochondrial rRNA. Uncleaved fragments were then enriched by 13 cycles of PCR before a final library purification using AMPure XP beads. Libraries were quantified with the Qubit dsDNA HS assay and assessed for quality and size distribution of library fragments using the Fragment Analyser and the NGS Fragment Kit (#DNF-473-0500).

#### DNA library preparation

Libraries were prepared from up to 300ng of each DNA sample using the Sure Select XT Target Enrichment for Illumina System according to the provided protocol for FFPE samples. DNA samples were sheared to an average fragment size of 150-200bp using the Covaris E220 Evolution and purified with Agencourt AMPure XP beads (Beckman Coulter Inc, #A6881). Purified DNA fragments were end-repaired to remove 3’ and 5’ overhangs before further purification with AMPure XP beads. A single ’A’ nucleotide was added to the 3’ ends of the blunt fragments to prevent them from ligating to another during the subsequent adapter ligation reaction, and a corresponding single ’T’ nucleotide on the 3’ end of the adapter provided a complementary overhang for ligating the adapter to the fragment. dA-tailed DNA was again purified with AMPure XP beads before paired-end adapters were ligated to the ends of the dA-tailed DNA fragments to prepare them for hybridisation onto a flow cell. After purification with AMPure XP beads 13 cycles of PCR were used to selectively enrich those DNA fragments that had adapter molecules on both ends and amplify the amount of DNA in the library suitable for target enrichment. Amplified gDNA libraries were purified using AMPure XP beads. gDNA libraries were quantified by fluorometry using the Qubit dsDNA HS assay and assessed for quality and fragment size using the Agilent Bioanalyser with the DNA HS Kit (#5067-4626). 750ng of each gDNA library (where available) was hybridised with target-specific probes (Canine All-Exon) before target molecules were captured on streptavidin beads and non-target molecules removed with a series of washes. Individual hybridised and captured libraries were then amplified for 12 cycles of PCR with unique indexing primers to allow multiplexed sequencing and purified with AMPure XP beads. Indexed libraries were quantified by fluorometry using the Qubit dsDNA HS assay and assessed for quality and fragment size using the Agilent Bioanalyser with the DNA HS Kit (#5067-4626). Quantification and fragment size data were used to calculate molarity for sequencing.

#### Sequencing

Sequencing was performed on the NextSeq 2000 platform (Illumina Inc, #20038897) using NextSeq 2000 P3 Reagents (200 Cycles) (#20040560). Libraries were combined in equimolar pools based on Qubit and Bioanalyser assay results and each pool was sequenced on a P3 flow cell. For RNA sequencing, PhiX Control v3 (Illumina Inc, #FC-110-3001) was spiked into each run at a concentration of 1% to allow troubleshooting in the event of any issues. Basecall data produced by the NextSeq 1000/2000 Control Software (Version 1.4.1.39716) was automatically converted into FASTQ files and uploaded to BaseSpace.

### Immunohistochemistry

#### Human OMM

Immunohistochemistry was conducted on 3µm-thick formalin-fixed paraffin-embedded human OMM tissue using a Leica Bond immunostainer (Leica Bond III Autostainer, Leica, #21.2201). A standard protocol was followed in accordance with the manufacturer’s instructions. CD3 was used to identify T cells and NK cells (Novocastra Liquid Mouse Monoclonal Antibody CD3, Leica Biosystems, #NCL-L-CD3-565), CD19 was employed to stain B lymphocytes, (Novocastra Liquid Mouse Monoclonal Antibody CD19, Leica Biosystems #NCL-L-CD19-163), CD68 was used to label monocytes/macrophages (Monoclonal Mouse Anti-Human CD68, Clone PG-M1, Dako, #M0876). Tissue was also stained with CD163 (CD163 Monoclonal Antibody, ThermoFisher Scientific #MA5-11458) CTLA4 (1:50 CTLA4 Polyclonal Antibody, Invitrogen, #PA5-115060) and cMET (1:400 HGFR/c-MET Antibody, 1G7NB, Novus Biologicals #44306SS) using the same protocol.

#### Canine OMM

Immunohistochemistry was conducted on 4µm-thick formalin-fixed paraffin-embedded canine OMM tissue, acquired using a microtome (Thermo Scientific Rotary Microtome Microm HM 340 E). All tissue sections, including relevant controls, were deparaffinized in xylene and rehydrated in graded ethanol through to distilled H20 prior to staining. Heat-induced antigen retrieval was performed using sodium citrate (pH 6.0; 110°C; 5-12 minutes overall) in a microwave (HistoS5, Milestone). Once retrieved, all tissue sections were stained in relevant batches in an autostainer (Epredia Autostainer 360) following a standard protocol which incorporates a 30-minute incubation with the primary antibody at room temperature. CD3 was used to identify T cells and NK cells (Novocastra Liquid Mouse Monoclonal Antibody CD3, Leica Biosystems, #NCL-L-CD3-565; 1/200 dilution), PAX5 was employed to stain B lymphocytes (Mouse Monoclonal Antibody Pax5, Becton and Dickinson, P67320-050; 1/50 dilution), and anti-IBA1 was used to label monocytes/macrophages (Rabbit polyclonal anti Iba1, Wako, 019-19741; 1/500 dilution). All sections were treated with Dako Real Peroxidase Blocking solution (Dako S2023) to block against endogenous peroxidase. The next step was a 40-minute incubation with either EnVision anti-mouse (Dako, EnVision+ System –HRP Labelled Polymer; 302059EFG_00) or Envision Anti-Rabbit. This system is based on an HRP labelled polymer which is conjugated with secondary antibodies. The labelled polymer does not contain avidin or biotin. As such, nonspecific staining resulting from endogenous avidin-biotin activity is eliminated or significantly reduced. Envision mouse was used to stain tissues with CD3 and PAX5, whereas EnVision Anti-Rabbit with DAB enhancer protocol was used for IBA1 (Dako EnVision®+ Dual Link System-HRP (DAB+), PD04048_02/K4065). All sections were then counterstained with Harris haematoxylin (Varistain Gemini Slide Stainer, Thermo Fischer Scientific), dehydrated with graded ethanol through to xylene, and covered with coverslips. Canine tissue was stained with CTLA4 and cMET using the same immunohistochemical protocol as in human OMM.

### Image acquisition

Slides were digitally scanned (Nanozoomer-XR, Hamamtsu Photonics K.K, Japan) and raw image data were saved in .ndpi format and handled by the software ndp.view 2 (Hamamatsu Photonics) to save details of the whole image in .jpeg format. Images were imported to ImageJ (FiJi Project, 2.11.0, GPLv3+) for labelling, and uploaded to QuPath (v0.5.0)[28].

### 16s sequencing

DNA was isolated, purified and quantified from FFPE core tissue samples from human and canine OMM as previously described. DNA was similarly isolated, purified and quantified from 4 FFPE core samples of normal canine oral tissue. Libraries were prepared from 2μL of each purified DNA sample using the Quick-16S™ Plus NGS Library Prep Kit (V3-V4, UDI) (Zymo Research, ∼D6421-PS1 and ∼D6421-PS2) according to the manufacturer’s protocol. Alongside the experimental samples, control libraries were prepared from the ZymoBIOMICS™ Microbial Community DNA Standard (positive), a ‘blank’ sample taken through the DNA extraction and purification processes without tissue (negative) and nuclease-free water (non-template control). The source material for both the positive control and NTC were provided in the Quick-16S™ Plus kit. The pool was purified using AMPure XP beads (Beckman Coulter, #A63881) to remove primer-dimer sequences. The pool was quantified by fluorometry using the Qubit dsDNA High Sensitivity assay (ThermoFisher Scientific, #Q32851) and assessed for quality and fragment size using the Agilent Bioanalyser with the DNA High Sensitivity Kit (Agilent #5067-4626). Fragment size and quantity measurements were used to calculate molarity for sequencing.

Sequencing (2x300) was performed on the NextSeq 2000 platform (Illumina Inc, #SY-415-1002) using the NextSeq 1000/2000 P1 Reagents (600 cycles) v3 Kit (#20075294). Loading concentration was 750pM as recommended in the Quick-16S Plus Kit User Guide. PhiX Control v3 (#FC-110-3001) library was spiked in at a concentration of 40% to help with cluster resolution and facilitate troubleshooting in case of any problems with the run.

## QUANTIFICATION AND STATISTICAL ANALYSIS

### Expression data analysis pipeline

#### Alignment and Gene-Level Counts

RNA-seq data were processed using the nf-core ’rnaseq’ pipeline v3.8.1[56–70]. In brief, samples were aligned via STAR v2.6.1d, and gene-based counts produced using Salmon v.1.5.2. Canine data were aligned to the ROS_Cfam_1.0 reference genome, annotated using the corresponding GTF file for build accession GCA_014441545.1. Human sequence data were aligned to hg38 and annotated with GENCODE.v41[71].

#### Unsupervised Clustering

Unsupervised consensus clustering of expression data was performed using the R Bioconductor package, ’cola’ [72]. Prior to clustering, gene level count data were subject to a variance stabilising transformation using DESeq2 [67], and the resulting matrix was used as an input. Five different clustering algorithms were tested, evaluating two to six clusters in each case. The cola algorithm resamples count data a fixed number of times, repeating the clustering process on each iteration. The optimum clustering strategy was selected on the stability of the resulting clusters; the method by which samples cluster most consistently.

#### Differential Expression Analysis

Differential expression analysis was performed using DESeq2. Models were fitted treating the clusters identified by cola, sex, and, where appropriate, batch as factors. Log fold change (logFC) estimates were produced using the apeglm shrinkage method [73], which is intended provide more robust estimates in the event of high within-group variability. Shrunken logFC estimates were accompanied by s-values [74], an aggregate false sign rate which are broadly analogous to q-values. P-values were also computed and adjusted using the IHW method [75]. Results were annotated using biomaRt [76], and volcano plots generated using EnhancedVolcano [77].

#### Over-Representation Analysis

Over-representation analysis (ORA) was performed via the R package ClusterProfiler[78]. ORA was performed for up-and down-regulated genes independently, taking the top differentially expressed genes with an s-value < 0.01 in each case. These gene lists were then tested for over-representation against the Biological Process (BP), Cellular Component (CC), and Molecular Function GO ontologies [79], and in addition to known pathways from the Kyoto Encyclopedia of Genes and Genomes (KEGG) [80]. Minimum and maximum gene set sizes were set to 10 and 1,000, respectively for all analyses.

### Immunocyte deconvolution

The computational framework CIBERSORTx was used to deconvolute the tumour-infiltrating immune cells from bulk RNA-sequencing data[27]. Twenty-two immune cell subtypes were parsed from the annotated gene signature matrix LM22 and 100 permutations of the CIBERSORTx web portal[27]. After running, only samples with CIBERSORT *p*-values < 0.05 were included in subsequent analyses. B-mode batch correction was applied, and quantile normalisation was disabled to generate absolute scores. CIBERSORT fractions for native B cells, memory B cells and plasma cells were aggregated into B cells; monocytes, M0, M1, M2 macrophages into monocytes/macrophages; and CD8, CD4, T follicular helper cells, regulatory T cells, and gamma delta T cells into T cells; and resting and activated NK cells into NK cells. Wilcoxon tests were used to compare the immune cell fractions between species, metastatic status, and transcriptomic consensus clustering groups. A cox-regression model was used to describe the time-to-event outcomes, with the adjustedCurves package [81] used to show the effect of a continuous variable adjusted for age and gender/sex. Receiver Operator Curves were plotted and optimal cut off points were defined using Youden’s index. Statistical significance was set at p<0.05.

### Image data analysis

Using the H&E slides to identify the tumour margins, the tumour areas were defined in QuPath. Positive cell detection was performed using the default parameters, except for sigma, which was set at 1.0μm. Wilcoxon tests were used to compare the ‘number of positive cells detected per mm^2^’ between transcriptomic consensus clustering groups. Optimal cut off points were defined as above. Statistical significance was set at p<0.05.

### 16s analysis

Cutadapt (v.4.5) was used to remove primers from paired-end 16S rRNA gene reads [82]. Both paired end reads were removed from further analyses if one or both of the pairs did not contain the 16S primers, using the –discard-untrimmed option. Mothur (v.1.48.0) was used for quality control, taxonomic assignment and out (operational taxonomic unit) clustering [83], following an adjusted version of the mothur Miseq pipeline [84]. Sequences were removed if they were >626bp in length, <322bp in length, contained ambiguous bases, had homopolymers of >9 bp in length, did not align to the correct variable region of the 16S rRNA gene or did not originate from bacteria. Chimeras were identified via the vsearch command, then removed. Sequence alignment and taxonomic assignment were conducted using the SILVA database [85] (v.138.1) formatted for use in mothur. OTUs were clustered using the “cluster.split” command in mothur. The OTU file, phylogenetic tree file, taxonomy file, and sample metadata file were packaged into a phyloseq object [86]

Data were decontaminated using decontam (v1.22.0)[87],whereby contaminants were identified by comparing the prevalence (presence/absence across samples) of each sequence feature in the true positive samples with the prevalence in negative controls. The threshold of 0.5 was established to designate as contaminants any exhibiting a higher prevalence in negative controls than positive controls. All contaminant sequence features were then removed from the entire dataset. Thirty-four contaminants were removed from the dataset, resulting in 33,929 operational taxonomic units (OTUs). Samples with <3000 reads were discarded. Two human and one canine sample did not have >3000 reads and were also discarded, resulting in 11 human OMM samples, 35 canine OMM samples, and 4 canine normal oral tissue samples.

To adjust for sequencing depth across samples, raw counts for each OTU were converted into relative abundance values by dividing by the total amount of counts for each sample. Goods coverage was calculated using phyloseq-coverage[86], and samples with a coverage value below 0.9 were considered to have insufficient sequencing depth and were discarded. To explore the diversity of microbial community composition within samples, we generated alpha-diversity indices using both Shannon diversity and Chao1 richness. Differences in alpha-diversity between cohorts were assessed by applying 2-sided Wilcoxon’s rank-sum tests. P values obtained from pairwise correlations were adjusted using the Benjamini-Hochberg procedure to control the false discovery rate. We presented significant findings as box and whisker plots.

To evaluate whether samples clustered by group according to their overall microbiota compositions, we conducted a beta-diversity analysis using PERMANOVA (Permutational Multivariate Analysis of Variance). This analysis was conducted with the *adonis2* function and Bray-Curtis dissimilarity matrix, implemented in the vegan package in R [88](version 2.6-4). We presented significant findings as principal component analysis (PCA) plots. Statistical significance was defined as p<0.05.

To determine the OTUs most likely to explain differences between groups, we conducted LefSE (Linear discriminant analysis effect size) analyses on variables exhibiting statistically significant alpha-or beta-diversity results. We used the R package, microbiomeMarker (1.8.0)[89], to identify markers generated using all phylogenetic levels, and presented results as tables and histographs.

## Supporting information

Supplementary Items

## Acknowledgements

We extend our heartfelt gratitude to all the patients or caregivers who generously consented to contribute samples for this study. This work was funded by the Kennel Club Charitable Trust (10915550_10917251); Wellcome Trust Institutional Translational Partnership Award (10587393_10587408); the Medical Research Council (MC_UU_00035/13), Melanoma Research Alliance and Rosetrees Trust (MRA Awards 687306, 917226), and supported by the Cancer Research UK Scotland Centre (CTRQQR-2021\100006). The authors thank the following colleagues for their expertise and assistance in this work: Jana Travnickova (University of Edinburgh), Vishad Patel and Craig Marshall (NHS Lothian BioResources), Jayne Hope and Cristina Vrettou (Roslin Institute), Steve Brawley (Vets4Pets), Lisa-Marie Butt (Edinburgh Bioquarter), and Scott Maxwell (Veterinary Pathology Unit, R(D)SVS), Helen Caldwell (University of Edinburgh), and Craig Nichol (University of Edinburgh). This manuscript was edited at Life Science Editors.

## Statement of author contributions

KLBB and EEP conceptualised and designed the study. Data collection was performed by KLBB, JdP, GP, LS, JT, DK, MP, JM, IB, SZ, SMG, DS, MT, DM, AM, KP, and MC. Data analysis was conducted by KD, YL, DS, AM, EEP, and KLBB. Data interpretation was contributed by LG, MS, MEM, KD, YL, KLBB, and EEP. KLBB and EEP conducted the literature search and generated the figures. All authors were involved in writing the paper and had final approval of the submitted and published versions. KLBB and EEP provided supervision and project administration.

