## Supplementary Items for "Oronasal mucosal melanoma is defined by two transcriptional subtypes in humans and dogs with implications for diagnosis and therapy"

**Bowlt Blacklock et al., Diagnostic sub-classification of oronasal mucosal melanoma**

**Supplemental Information Items**

**Supplemental Figure 1:** Clinical data and Kaplan Meier survival plots associated with human (Edinburgh) and canine (Bowlt Blacklock) OMM cohorts

**Supplemental Figure 2:** Kaplan-Meier survival plots and COSMIC signature data associated with human and canine OMM

**Supplemental Figure 3:** Two shared transcriptomic subgroups stratify OMM in human and canine patients

**Supplemental Figure 4:** Violin plots, ROC curves, and Kaplan Meier survival plots associated with transcriptomic subgroup

**Supplemental Data 1:** Pathway analysis results associated with two transcriptomic subtypes identified in human and canine OMM.

**Supplemental Data 2**: List of 812 human and canine homologous genes inputted into the randomForest machine learning algorithm and the genes utilised by the model.

**Supplemental Data 3**: Immunocyte infiltration in human and canine OMM, parsed from the annotated gene signature matrix LM22 and 100 permutations of the CIBERSORTx web portal.

**Supplemental Data 4:** Beta diversity metrics for microbiome data from human and canine OMM.

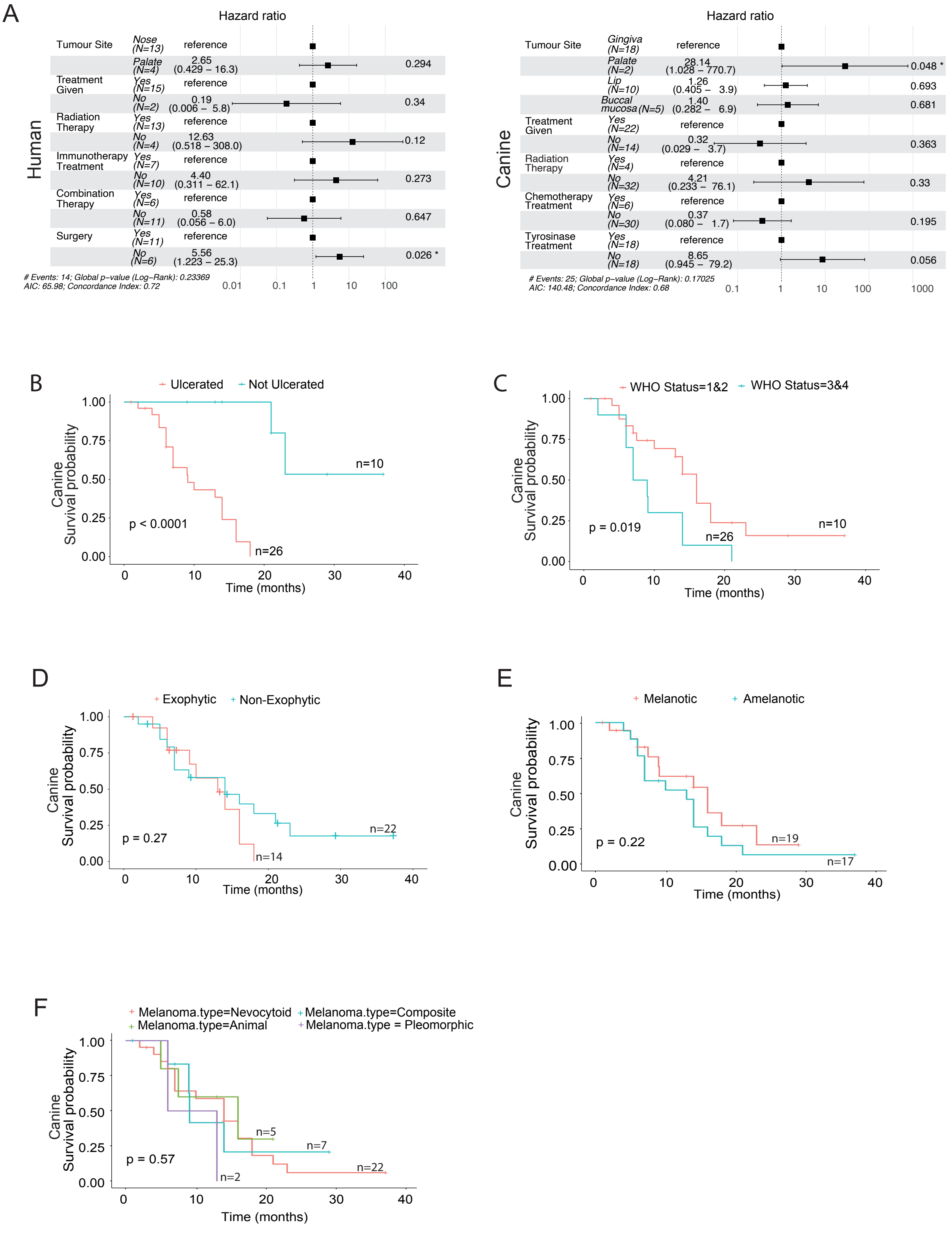
A
Hazard ratio
Human
Tumour Site
Nose (N=13) reference
Palate (N=4) 2.65 (0.429 – 16.3) 0.294
Treatment Given
Yes (N=15) reference
No (N=2) 0.19 (0.006 – 5.8) 0.34
Radiation Therapy
Yes (N=13) reference
No (N=4) 12.63 (0.518 – 308.0) 0.12
Immunotherapy Treatment
Yes (N=7) reference
No (N=10) 4.40 (0.311 – 62.1) 0.273
Combination Therapy
Yes (N=6) reference
No (N=11) 0.58 (0.056 – 6.0) 0.647
Surgery
Yes (N=11) reference
No (N=6) 5.56 (1.223 – 25.3) 0.026 *
### Events: 14; Global p-value (Log-Rank): 0.23369
AIC: 65.98; Concordance Index: 0.72
0.01 0.1 1 10 100
Hazard ratio
Canine
Tumour Site
Gingiva (N=18) reference
Palate (N=2) 28.14 (1.028 – 770.7) 0.048 *
Lip (N=10) 1.26 (0.405 – 3.9) 0.693
Buccal mucosa (N=5) 1.40 (0.282 – 6.9) 0.681
Treatment Given
Yes (N=22) reference
No (N=14) 0.32 (0.029 – 3.7) 0.363
Radiation Therapy
Yes (N=4) reference
No (N=32) 4.21 (0.233 – 76.1) 0.33
Chemotherapy Treatment
Yes (N=6) reference
No (N=30) 0.37 (0.080 – 1.7) 0.195
Tyrosinase Treatment
Yes (N=18) reference
No (N=18) 8.65 (0.945 – 79.2) 0.056
### Events: 25; Global p-value (Log-Rank): 0.17025
AIC: 140.48; Concordance Index: 0.68
0.1 1 10 100 1000
B
Ulcerated
Not Ulcerated
Canine Survival probability
n=10
n=26
p < 0.0001
Time (months)
C
WHO Status=1&2
WHO Status=3&4
Canine Survival probability
n=10
n=26
p = 0.019
Time (months)
D
Exophytic
Non-Exophytic
Canine Survival probability
n=22
n=14
p = 0.27
Time (months)
E
Melanotic
Amelanotic
Canine Survival probability
n=19
n=17
p = 0.22
Time (months)
F
Melanoma.type=Nevocytoid
Melanoma.type=Composite
Melanoma.type=Animal
Melanoma.type = Pleomorphic
Canine Survival probability
n=5
n=7
n=22
n=2
p = 0.57
Time (months)

**Supplementary Figure 1: Clinical data and Kaplan Meier survival plots associated with human (Edinburgh) and canine (Bowlt Blacklock) OMM cohorts**

**A:** Hazard ratios for survival in human (left) and canine (right) patients with OMM. **B-F:** Kaplan-Meier survival plots for canine patients based on (**B**) tumour ulceration**,** (**C**) patient WHO status, (**D**) tumour exophytic status**, (E)** tumour melanotic status, and (**F**) tumour histopathological subtype **(F).** The y-axis represents survival probability and the x-axis denotes time, with tick marks indicating censored data points. A similar comparison in humans was not possible.

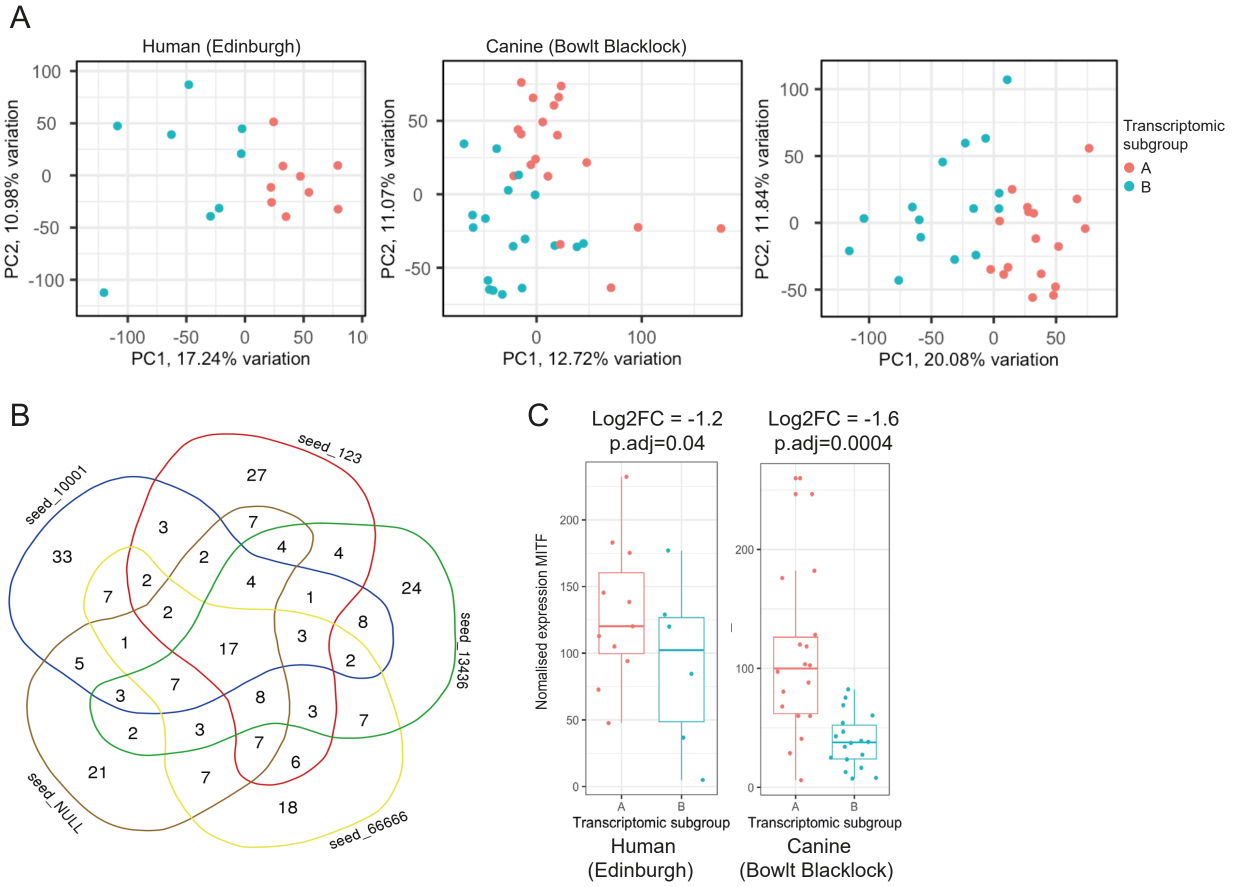
A
Human (Edinburgh)
Canine (Bowlt Blacklock)
PC1, 17.24% variation
PC2, 10.98% variation
PC1, 12.72% variation
PC2, 11.07% variation
PC1, 20.08% variation
PC2, 11.84% variation
Transcriptomic subgroup
A
B
B
seed_10001
seed_123
seed_13436
seed_66666
seed_NULL
C
Log2FC = -1.2
p.adj=0.04
Log2FC = -1.6
p.adj=0.0004
Nomalised expression MITF
Transcriptomic subgroup
Transcriptomic subgroup
Human
(Edinburgh)
Canine
(Bowlt Blacklock)

**Supplementary Figure 2: Two shared transcriptomic subgroups stratify OMM in human and canine patients**

**A:** Principal component analysis (PCA) plots for human (Edinburgh)(n=17), canine (Bowlt Blacklock) (n=36), and canine (Prouteau)(n=32) OMM, with cells coloured by transcriptomic subgroup. **B:** Venn diagram of 5 models showing 41 genes consistently ranked among the top 100 genes shared by at least 4 models and 17 genes shared by all five models. **C:** Box and whisker plots showing MITF expression levels in human and canine OMM according to transcriptomic subgroup

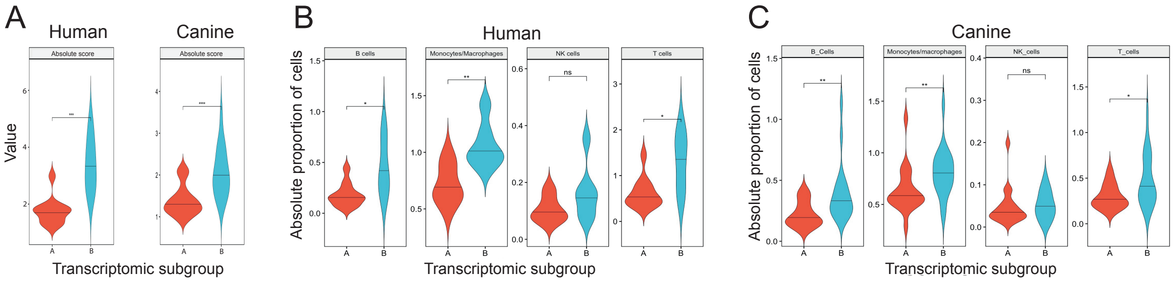


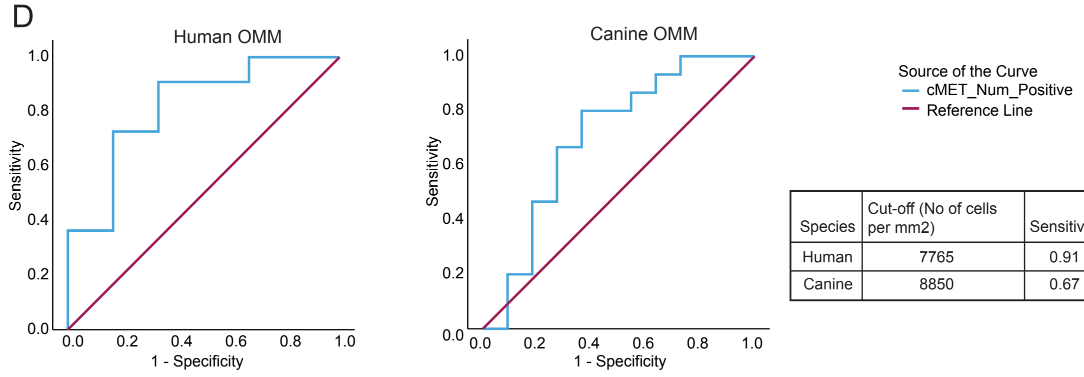


| Species | Cut-off (No of cells per mm2) | Sensitivity | Specificity |
|---|---|---|---|
| Human | 7765 | 0.91 | 0.67 |
| Canine | 8850 | 0.67 | 0.82 |

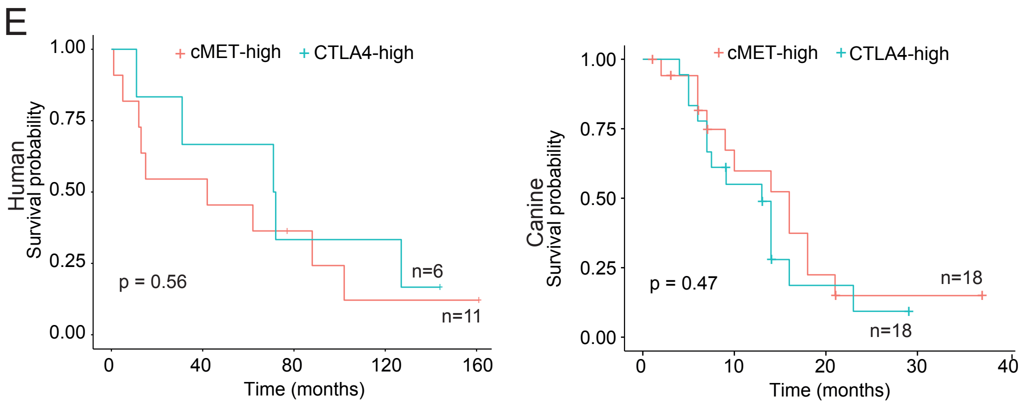

**Supplementary Figure 3: Violin plots, ROC curves, and Kaplan Meier survival plots associated with transcriptomic subgroup**

**A**: Violin plots of CIBERSORT "Absolute Score", representing total immune cell infiltration, in human and canine OMM. **B, C:** Violin plots showing the absolute proportion of immune cell types in humans and canine OMM. **D:** ROC curves and optimal cut-off parameters for number of cMET-positive cells per $mm^2$ in human and canine OMM. **E:** Kaplan-Meier plot showing survival stratified by transcriptomic subgroup in human and canine patients with OMM.
